# A Survey on Optimization and Machine Learning-Based Fair Decision Making in Healthcare

**DOI:** 10.1101/2024.03.16.24304403

**Authors:** Zequn Chen, Wesley J. Marrero

## Abstract

The unintended biases introduced by optimization and machine learning (ML) models are a topic of great interest to medical researchers and professionals. Bias in healthcare decisions can cause patients from vulnerable populations (e.g., racially minoritized, low-income, or living in rural areas) to have lower access to resources and inferior outcomes, thus exacerbating societal unfairness. In this systematic literature review, we present a structured overview of the literature regarding fair decision making in healthcare until April 2024. After screening 801 unique references, we identified 114 articles within the scope of our review. In our review, we comprehensively examine fair decision-making methodologies in healthcare by systematically identifying and categorizing biases within both data and models. Initially, we elucidate existing bias within healthcare decision making. Then, we present a range of fairness metrics drawn from different use cases, followed by analyzing and classifying bias mitigation strategies into pre-processing, in-processing, and post-processing techniques. We provide a broad conceptual overview and practical illustrations of each approach. Additionally, we examine emerging bias mitigation technologies that, though not yet applied in healthcare, show substantial promise for future integration. Our review aims to increase awareness of fairness in healthcare decision making and facilitate the selection of appropriate approaches under varying scenarios.

## 1. Introduction

An increasing number of healthcare researchers and practitioners are leveraging quantitative methodologies to improve decision making. These methodologies aim to achieve desirable resource allocation strategies, testing procedures, or treatment protocols. Optimization and machine learning (ML) models and algorithms have been proven to be extremely helpful in medical decision making and health policy [1–7]. For example, researchers have been applying optimization models to schedule patients based on their predicted no-show rates; and reinforcement learning has been used for treatment recommendations to ensure effective prescription and low mortality rates [8, 9]. However, such advanced decision-making methods can lead to inequitable outcomes as they may not give sufficient attention to underrepresented groups [10], such as those who are racially minoritized or low-income. Our survey presents a review of fair decision-making techniques, revealing how we can leverage optimization and ML approaches to ensure equitable access to healthcare.

The unintended biases introduced by optimization and ML are of special interest to practitioners and researchers [11–14]. For example, compared to White veterans, the medical policies generated by reinforcement learning often offer Black veterans fewer opportunities to receive cardiovascular screenings. Hispanic patients are disproportionately underdiagnosed by convolutional neural networks because they tend to have limited access to healthcare resources and the ML approach cannot perform satisfyingly with inadequate data [15]. Clinicians have utilized ML models in Warfarin dosing, which shows superior performance in European patients but unsatisfying outcomes in Asian patients [16, 17]. Such bias may cause decision makers to distribute fewer medical resources to racially minoritized subgroups.

### 1.1 General Trends in Fair Decision Making

Recent research in ML and optimization-based decision making in healthcare has increasingly centered on three complementary methodological paradigms: pre-processing, in-processing, and post-processing. *Pre-processing* techniques focus on transforming and rebalancing raw data before model training to mitigate historical biases and improve representation for underrepresented groups. For instance, methods such as fair data transformers and advanced natural language processing pipelines are being developed to ensure that the input data itself is less biased. *In-processing* methods embed fairness directly into the learning or optimization process by incorporating fairness penalties, constraints, or fairness considerations in the algorithm or model design. These methods include the adaptation of fair reward design in reinforcement learning, as well as formulation of optimization problems (e.g., mixed-integer or stochastic programming models) that inherently account for equitable decision criteria during model training. Finally, *post-processing* approaches adjust model outputs after training to correct any residual biases. Techniques like Laplacian smoothing and multi-accuracy adjustments are employed to refine decision policies, ensuring fair resource allocation and treatment recommendations across diverse patient populations. Collectively, these research avenues are advancing the state-of-the-art in achieving both high-performance and equitable decision making in healthcare.

### 1.2 Existing Systematic Reviews

Past literature reviews focus on the biases brought or perpetuated by ML in prediction settings. For example, Ahmad and collaborators show that ML-based predictions often yield fewer satisfying outcomes in underrepresented groups due to their insufficient data [18]. Mishler and coauthors reveal that predictors are sensitive to proportions of different populations, and incorporating fairness definitions can help avoid such issues [19]. In contrast to previous reviews centering on prediction, we focus on fairness within the context of decision making. It is worth noting that our review has some overlaps with Smith et al.’s survey [14]. The main differences between our surveys are: 1) their work focuses on fairness in reinforcement learning exclusively, while our paper explores other in-processing techniques such as mixed-integer programming (MIP) and stochastic programming; 2) we review different pre-processing techniques, such as fair data transformers and natural language processing; and 3) we consider post-processing methods such as Laplacian smoothing and multi-accuracy approaches. The fair reinforcement learning methods cited in Smith et al. are included in our review for completion purposes [20–23]. However, we refer the interested reader to their review for an in-depth description of these works.

### 1.3 Contributions

Our review provides a comprehensive overview of fair decision-making approaches in healthcare. Its main contributions are: (1) We systematically identify and categorize biases affecting both data and models, summarizing multiple fairness metrics (and their limitations) through concrete examples; (2) We analyze the advantages and disadvantages of existing bias-mitigation strategies for decision making in pre-processing, in-processing, and post-processing techniques. Even though several reviews have surveyed fairness methods within the context of prediction [24–26], our work aims to summarize the understanding of fairness for decision-making settings; (3) We examine outstanding challenges in fair decision making and proposing promising directions for future research; (4) We explore emerging bias-mitigation technologies that, while not yet applied in healthcare, offer significant potential for future integration; and (5) We clarify both high-level concepts and detailed examples of each methodology, thereby serving as a practical resource for both practitioners and researchers.

### 1.4 Organization of the Systematic Review

We begin by describing our literature search strategy. Then, we portray bias categories commonly encountered in decision making, followed by a summary of fairness metrics. Next, we present bias mitigation techniques to extend the traditional decision-making framework while achieving fair choices. Finally, we outline our survey’s contributions and limitations, how our survey can assist researchers and practitioners, as well as promising future directions.

## 2. Search Strategies

For our systematic review, we searched the Google Scholar database for records related to fair decision making in healthcare. The electronic search strategy used the terms “decision making” and “healthcare”, combined with a term in “bias”, “fairness”, or “equity”, and one of the terms in “optimization”, “machine learning”, “deep learning”, “reinforcement learning”, “game”, or “network.” The keyword combinations we used are demonstrated in Figure 1. We included all records mentioning these keywords in the publication title, abstract, or full text. The publication language was restricted to English. The search’s last update was in April of 2024.

**Figure 1:**
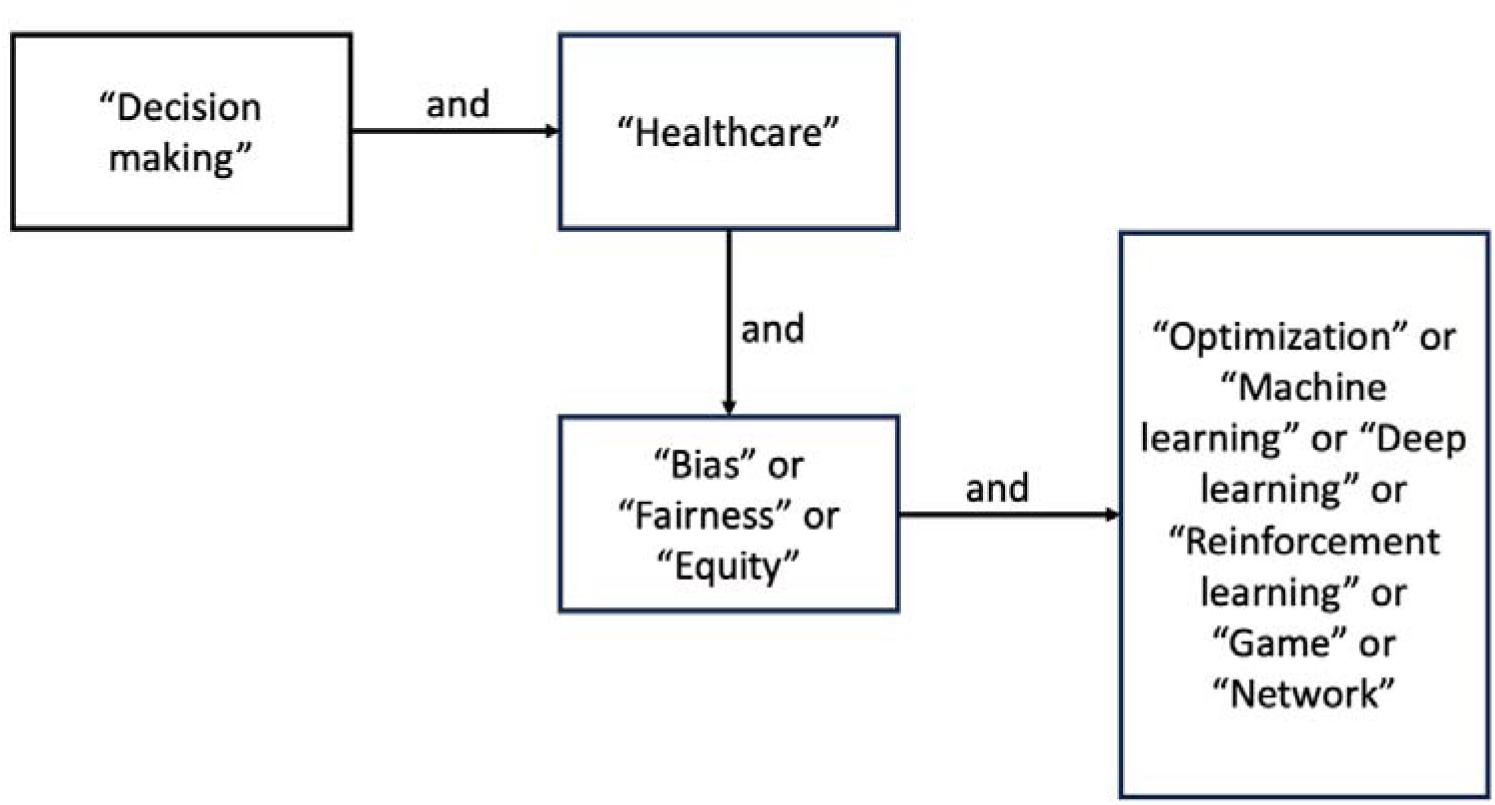
Key words combination for literature review.

The titles and abstracts of the articles were screened by one author referred to as “investigator,” then the selected manuscripts were double-checked by another author referred to as “researcher.” Records were excluded if the publications: 1) did not address healthcare topics and could not be extended to healthcare easily; 2) did not focus on decision making (e.g., papers focusing on predictions); or 3) only included introductory text or conference abstract. Of the remaining publications on methodology or review of bias, fairness metrics, and bias mitigation methods, the full text was screened by investigator before the final discussion with researcher. The articles and surveys are categorized into three sections: bias in healthcare (section 3.1), fairness metrics (section 3.2), and fair decision-making models and algorithms (section 3.3). Discordance between authors was settled through discussion until the consensus had been achieved.

Fair decision-making concepts (i.e., types of biases, fairness metrics, or bias mitigation approaches) were extracted from the full text of the selected works by investigator. To ensure the accuracy and completeness of the extracted aspects, the selected concepts were double-checked by researcher. All chosen concepts were grouped into section 3.1, section 3.2 or section 3.3 to reflect their relations.

## 3. Literature Review Results

The systematic review led to 801 records; 211 of them were unrelated and could not be easily transferred to the healthcare domain, and 212 of them did not focus on decision making. Furthermore, 72 records were excluded because they were introductory text or conference abstracts. Of the remaining articles, 155 papers discussed fair decision making in healthcare without a focus on methodology or specific examples of application. In the 151 papers left, we excluded 37 additional articles since they did not meet the inclusion criteria described in section 2. Of the remaining 114 papers, 25 were review papers and 89 were original papers. The flow diagram for literature review is shown in Figure 2.

**Figure 2:**
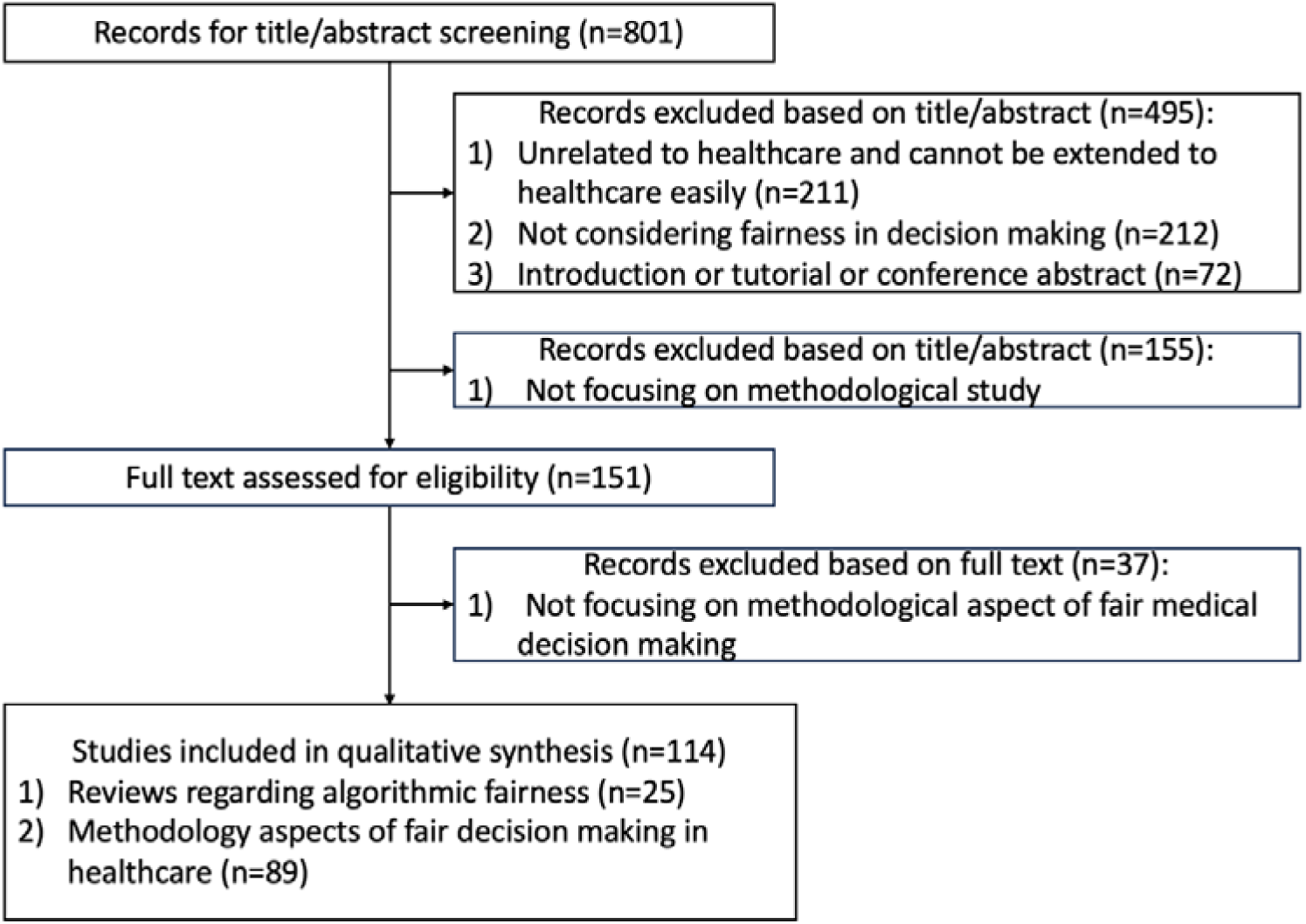
Flow diagram for the systematic literature review of fair decision making in healthcare.

Of the included papers, all concepts related to fair decision-making in healthcare were extracted. We created a structured coverage of the key findings in the following sections. section 3.1 categorizes biases into three groups: 1) algorithmic bias, 2) data bias, and 3) publication bias. section 3.2 classifies fairness metrics as one of the following three types: 1) fairness through unawareness, 2) demographic parity, and 3) equal opportunity. section 3.3 categorizes bias mitigation techniques into three distinct classes: pre-processing, in-processing, and post-processing methodologies.

For the in-processing section, we describe the cutting-edge optimization and ML methods used to alleviate bias. At a high level, there are two mainstream approaches to achieving fairness in these methods: incorporating fairness in objectives or adding fairness-enhancing constraints.

### 3.1 Bias in Healthcare Decision Making

Biases can exist in data and algorithmic design, which may impede decision-making systems from generating equitable outcomes among subgroups. In this section, we summarize some sources of bias impacting decision making across healthcare domains. These biases can be categorized into one of the following classes: algorithmic bias, data bias, and publication bias. section 3.3 of our survey will summarize methods addressing these biases.

#### 3.1.1 Algorithmic Bias

Algorithmic bias stems from computational procedures failing to consider fairness in their execution. This type of bias is a result of improper algorithmic design, which consequently influences fairness of medical decisions [27]. For example, optimization-based vaccine allocation algorithms aiming to maximize overall social welfare may exacerbate demographic disparities since underrepresented populations may have less access to vaccines under this objective [28]. Similarly, ambulance allocation models may fail to consider fairness of unit availability across different populations by solely maximizing the overall survival rate. While the survival rate may be high among the entire population, it can be low among patients from vulnerable populations, such as people with lower socioeconomic status [29]. Algorithmic bias also exists in ML models. For instance, Samorani and coauthors have found that ML-based scheduling models have a higher likelihood of assigning Black patients to overbooked slots [30], resulting in worse service experience and longer waiting time.

#### 3.1.2 Data Bias

Data bias refers to the unfairness generated by prejudiced data sources. Unbiased datasets are often necessary for high-quality decision-making models [31]. However, socioeconomic and racial disparities in resource availability may lead to skewed datasets [32]. Two common data biases in healthcare are aggregation biases and representation biases. Aggregation bias refers to the effect of aggregating data without considering disparities among subgroups [33]. For example, hemoglobin A1c level, a widely accepted indicator of diabetes, vary across sex and ethnicity. If we ignore the subgroup differences in the data and draw conclusions for subpopulations based on the entire population, we may introduce aggregation biases [10, 11, 34, 35]. Representation bias occurs when the data cannot represent the characteristics of all subgroups [36–40]. For instance, providers may have fewer electronic health records for people from lower socioeconomic status as they have limited access to electronic healthcare systems. Hence, we can expect fewer data among people from lower socioeconomic status, which indicates their data cannot demonstrate their overall characteristics. If decision-making models are built with data underrepresenting this population, the developed models can be biased against those with lower socioeconomic status [41, 42].

Another source of data bias is response bias. Response bias occurs when data are labeled inconsistently or collected by unreliable methods. Response bias frequently happens in self-reported data or surveys due to participants’ inaccurate answers. For instance, when medical students rate their mental health conditions in surveys, they tend to provide socially acceptable answers. However, such answers often do not align with their true mental conditions as students [43]. Since policymakers may harness data to make public health decisions, response bias can skew decision making [44], and models built from data with response bias may underestimate the seriousness of medical students’ mental health problems [32].

#### 3.1.3 Publication Bias

Publication bias happens when the researchers’ decision to publish a paper depends on the study results. Compared with studies without positive results, publishing medical studies with positive results is generally easier [45]. This phenomenon may lead to the overestimation of certain clinical treatments as evidence against a treatment is not made available. Medical practitioners may then make treatment decisions based on biased outcomes, giving rise to degraded treatment effects.

An example of publication bias occurred during the COVID-19 pandemic. The academic papers concerning COVID-19 treatment were published rapidly within this period. Most publications showed promising outcomes of COVID-19 treatments, while less satisfying research results only had a slim chance of being published [46]. When related treatments were applied to the general population, many of these treatments produced inferior outcomes compared to publication results [46]. Another example comes from the Cochrane Review on topical benzoyl peroxide, a widely used acne treatment. In 2019, Yang and coauthors found that most benzoyl peroxide studies before 2015 were still unpublished because of their negative results [47]. This finding suggests published literature could not reliably reflect the overall effect of benzoyl peroxide, and medical practitioners applying benzoyl peroxide may not have achieved the desired treatment effects.

### 3.2 Fairness Metrics

In this section, we present the evaluation of three types of metrics: fairness through unawareness, demographic parity and equal opportunity. To portray the ideas of different fairness metrics, we use a vaccine distribution example. For illustration purposes, we restrict our attention to only one binary sensitive attribute, denoted by A. An example of this type of attribute may be a dichotomized version of race, which includes White and people of color as its categories. The metrics covered by the section are summarized in Table 1.

**Table 1:** Definition of fairness metrics.

|  | **Fairness**<br>**through**<br>**unawareness** | **Demographic parity** | **Equal opportunity** |
| --- | --- | --- | --- |
| Sensitive<br>attribute | No | Yes | Yes |
| Individual<br>qualification | No | No | Yes |
| Definition | $X \cap g = \emptyset$ | After feeding $X$ to<br>decision-making models,<br>$E(r_{g_1}) = E(r_{g_2})$ | After feeding $X$ to<br>decision-making models,<br>within a qualified<br>subgroup,<br>$E(r_{g_1}) = E(r_{g_2})$ |

In this review, *x* denotes entire characteristics of the population, *g* denotes the sensitive attribute (in our case sensitive attribute is race), *r*_*g*_1__ denotes the cumulative rewards of White patients while *r*_*g*_2__ denotes the cumulative rewards of people of color. A qualified subgroup represents a subset of the general population that may be of special interest to decision makers. For example, certain patients, such as senior citizens and those residing in areas with inadequate healthcare resources, could be considered qualified patients as they may be more susceptible to the disease.

#### 3.2.1 Fairness Through Unawareness

Fairness through unawareness is the base fairness metric [15]. It does not consider any sensitive attribute during the decision-making process. Within the context of our vaccination distribution example, fairness through unawareness means the model is considered fair if it does not consider race while deciding the vaccination distribution. The benefit of fairness through unawareness is it is simple and convenient to implement. The advantage of fairness through unawareness lies in its simplicity and ease of implementation. However, simply ignoring sensitive attributes may not remove inequity, because other variables can be highly correlated with the sensitive traits. For instance, socioeconomic status or geographic location could be strongly correlated with race. Consequently, the model might still make decisions that indirectly disadvantage certain people of color, perpetuating inequity. This method has been proven to be invalid in many cases since the method neglects the structural inequalities embedded in proxy variables [48].

#### 3.2.2 Demographic Parity

The demographic parity fairness metric aims to ensure the expected cumulative reward of a decision-making model is independent of sensitive attributes [13]. Independence of sensitive attributes indicates the outcomes (i.e., expected cumulative rewards) must be equivalent in privileged and unprivileged groups [49]. In our vaccine distribution example, demographic parity ensures that, when other information is the same (e.g., age, socioeconomic status), White patients and patients of color have the same expected cumulative rewards (e.g., total symptom improvements). The problem with demographic parity is that it does not consider the population’s ground-truth qualifications. For instance, consider a case where patients of color are statistically more vulnerable to a particular disease due to underlying health disparities or systemic inequities. These patients might have a higher ground-truth qualification for receiving the vaccine. Applying demographic parity in such a scenario would require the vaccines be distributed equally across racial groups, regardless of the groups’ actual vulnerability to the disease. This distribution would neglect the real-world needs of patients of color and could lead to suboptimal outcomes, such as under-protection of the most vulnerable populations.

However, demographic parity may be appropriate in a scenario where there is no significant difference in vulnerability across racial groups, and fairness requires strict equality of treatment. For example, if the disease impacts White patients and patients of color equally and their ground-truth qualifications are the same (i.e., both groups are equally at risk and stand to benefit equally from the vaccine), demographic parity ensures that neither group is disadvantaged in the vaccine distribution process. In such a situation, applying demographic parity would help prevent systemic biases from skewing vaccine access while maintaining fairness in outcomes.

#### 3.2.3 Equal Opportunity

Similar to demographic parity, equal opportunity verifies whether the expected cumulative rewards for privileged and unprivileged groups are the same [27]. However, demographic parity applies to the entire population, while equal opportunity applies solely to a qualified population [49]. Within our example, this metric requires that within a qualified population, the cumulated expected reward of receiving vaccine among the unprivileged group (*A = people of color*) is the same as the cumulated expected reward of receiving vaccine among the privileged group (*A = White*).

Equal opportunity requires a well-defined notion of “true eligibility,” meaning that individuals who meet the criteria for a treatment should be treated equally across groups. Within our vaccine distribution example, if White and patients of color both have a set of equally high-risk individuals, equal opportunity can guarantee that no group is unfairly advantaged in the eligible, high risk, population.

Nevertheless, equal opportunity becomes inappropriate when unqualified individuals are unfairly impacted when the qualification criteria themselves are biased or imperfectly measured. For instance, if systemic issues lead to more misclassifications of people of color as unqualified, focusing exclusively on the qualified population may overlook these disparities and exacerbate existing inequities.

#### 3.2.4 Summary of Fairness Metrics

Fairness through unawareness is the simplest fairness metric, requiring that sensitive attributes, such as race or gender, are excluded from decision making. However, it is often invalid because other variables correlated with sensitive attributes can serve as proxies, perpetuating biases even when the sensitive attributes are omitted. Demographic parity, on the other hand, evaluates whether decision making is independent of sensitive attributes across the entire population, ensuring equality of outcomes. While this can help mitigate systemic biases, it ignores differences in ground-truth qualifications between groups, potentially leading to unfair decisions. Equal opportunity refines demographic parity by focusing on whether decision making is independent of sensitive attributes within a qualified subgroup, such as high-risk individuals in a vaccine distribution example. This approach ensures fairness among those most in need but does not address inequities among the unqualified population, such as those misclassified as ineligible or indirectly disadvantaged. These metrics vary in applicability, with their effectiveness depending on the context and the specific fairness goals.

### 3.3 Bias Mitigation

In this section, we summarize different bias mitigation approaches used across healthcare domains. The methods can be categorized into pre-processing, in-processing, and post-processing. Pre-processing mechanisms clean and manipulate the input data before it is used in decision-making models or algorithms [50]. In-processing methodologies refer to building unbiased models or algorithms directly [51]. Post-processing methods calibrate algorithmic outcomes to achieve fairness [52].

#### 3.3.1 Pre-Processing

Datasets may be biased, which can cause skewed decisions. For instance, when a dataset is imbalanced, decisions may be biased toward subpopulations with smaller sizes [53]. Pre-processing methods can help circumvent possible biases in this setting. There are five commonly used approaches for pre-processing: reweighting the underrepresented populations, resampling, fair data transformers, natural language processing, and post-survey analysis. The identified pre-processing methods, along with their respective reference, areas of application, fairness metrics, and targeted bias categories, are summarized in Table 2. Please see section 1.1 in the Supplementary Information for an in-depth discussion of the strengths and limitations of the pre-processing methods presented in this review.

**Table 2:**
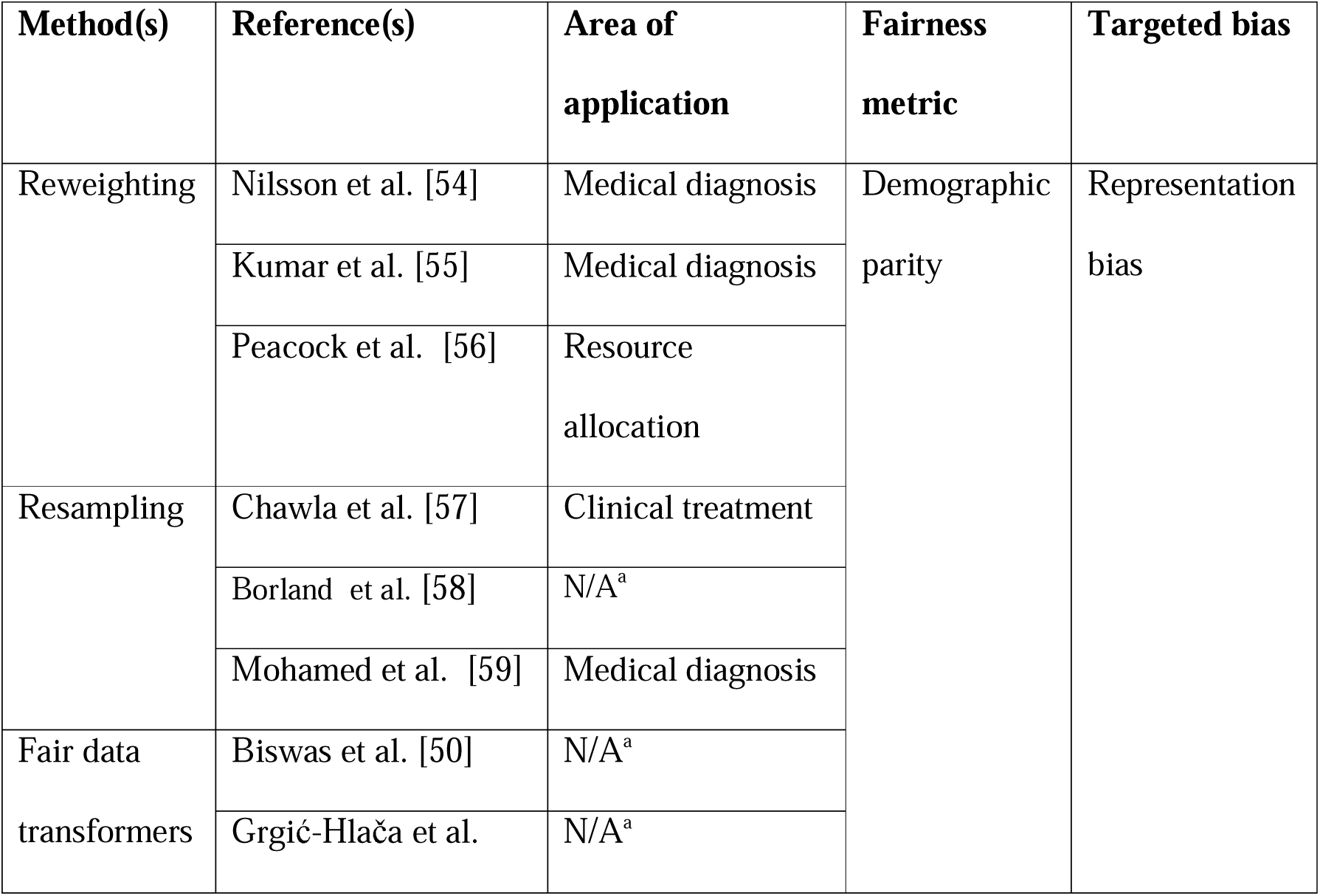
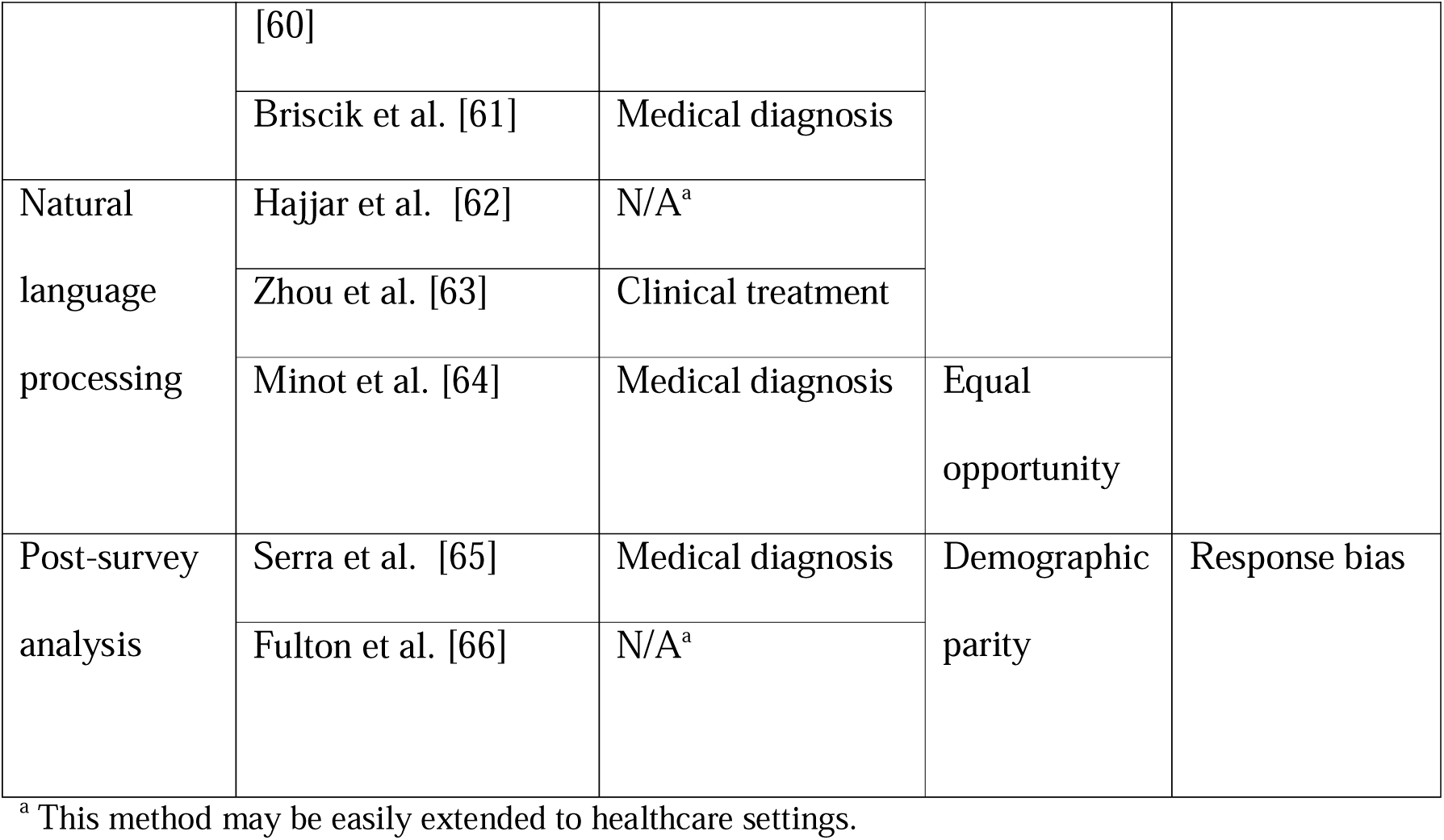
Pre-processing bias mitigation methods.

#### Reweighting

Reweighting assigns greater weights to underrepresented instances [54]. Biases may be introduced to decision-making tasks if we fail to process underrepresented population’s data. Skewed data can also lead to threatening consequences for underrepresented populations. For example, African Americans and Asians have fewer instances in genome studies, which gives rise to higher misclassification rates for the two subgroups in clinical research [11]. Classification methods can investigate the features of each piece of data and use them to decide which category or label the data belongs to. Since physicians may rely on classification methods for diagnosis and treatment design, varying misclassification rates for different subgroups can trigger bias in decision making. A remedy for similar issues is to assign greater weights to underrepresented instances, hence data from all populations play an equal role in the modeling process [54]. Kumar and coauthors have shown that distributing more weight to underrepresented groups can improve fairness in medical image classification by 8% [55].

One crucial factor influencing reweighting efficiency is the choice of weighting scheme. Complex or highly granular schemes (e.g., accounting for multiple intersecting protected attributes) can increase computational overhead. Another consideration is the distribution of protected subgroups: extremely small or imbalanced groups may lead to unstable or excessively large weights, thereby affecting convergence speed and model stability.

#### Resampling

Resampling is a technique to ensure the data is balanced (i.e., has an near-equal number of instances from each subgroup) by repeatedly drawing samples from the same data [53]. Using imbalanced data directly may favor the majority groups while disregarding the minority ones. To avoid this concern, we can resample from the minority groups, so the majority and minority groups have approximately the same size. The relative proportions of protected groups heavily influence efficiency of resampling [53]. When there is a severe imbalance between groups, oversampling the minority group or undersampling the majority group can lead to overfitting or information loss, respectively.

#### Fair data transformers

Fair data transformers extract features from the input data in a fair manner. Such transformers modify the input data to achieve fairness [50]. Data transformers, such as principal component analysis, are popular techniques for pre-processing data. In practice, multiple transformers are typically evaluated for a specific fairness metric (e.g., demographic parity). The transformers achieving the fairest output are used to produce inputs for ML and optimization algorithms. Biswas and collaborators have shown that a proper data transformer can significantly improve the fairness of outcomes [50]. Though fair data transformers have not been deployed in healthcare to the best of our knowledge, it is easy to extend a fair data transformer to healthcare data preprocessing. For example, in the context of vaccination distribution, we can collect data such as age, sex, population density, and incidence rate in the areas where individuals live. Then, we apply several fair data transformers and feed the transformed data into the same decision-making algorithm. After the algorithm outputs vaccination distribution decisions, we can evaluate the fairness of outcomes and choose the data transformer that produces the fairest output.

The efficacy of a fair data transformer is sensitive to several factors, including the quality and representativeness of patient data, the complexity of fairness definitions, and the regulatory environment in which it operates. For instance, imbalanced datasets or limited data on certain patient subgroups can reduce the transformer’s ability to produce equitable outcomes. Additionally, ensuring that the transformed features remain clinically interpretable is vital for practitioner trust, and any deployed system must be monitored over time to adapt to changing patient populations and medical standards.

#### Natural language processing

Natural language processing removes biased information from text data before feeding data to decision-making algorithms [63]. Due to the complexity of healthcare data, natural language processing is becoming increasingly popular in fair pre-processing settings.

#### Post-survey analysis

Post-survey analysis for data bias mitigation refers to the process of analyzing survey data after its collection to identify, assess, and correct various types of biases that may have been introduced during the data collection phase [31]. For example, some respondents might misremember their health history; hence the treatment effect for them is likely to be inferior compared to respondents remembering correctly. Researchers can mitigate this issue by cross-referencing survey responses with medical records to mitigate such bias [65].

#### 3.3.2 In-Processing

In-processing methodologies incorporate one or more fairness metrics directly in the design of algorithms or models to lessen biases. These bias mitigation techniques are attracting increased attention within domains such as resource allocation, scheduling, and clinical treatment [30, 67]. Overall, we find six typically used in-processing methods in the literature: mixed-integer programming, stochastic programming, deep reinforcement learning, fair survival analysis, multi-objective Markov Decision Process, and constrained Markov Decision Process. Table 3 demonstrates the methods and the corresponding references and applications. Please refer to section 1.2 of the Supplementary Information for a detailed explanation of the strengths and limitations of in-processing methods.

**Table 3:**
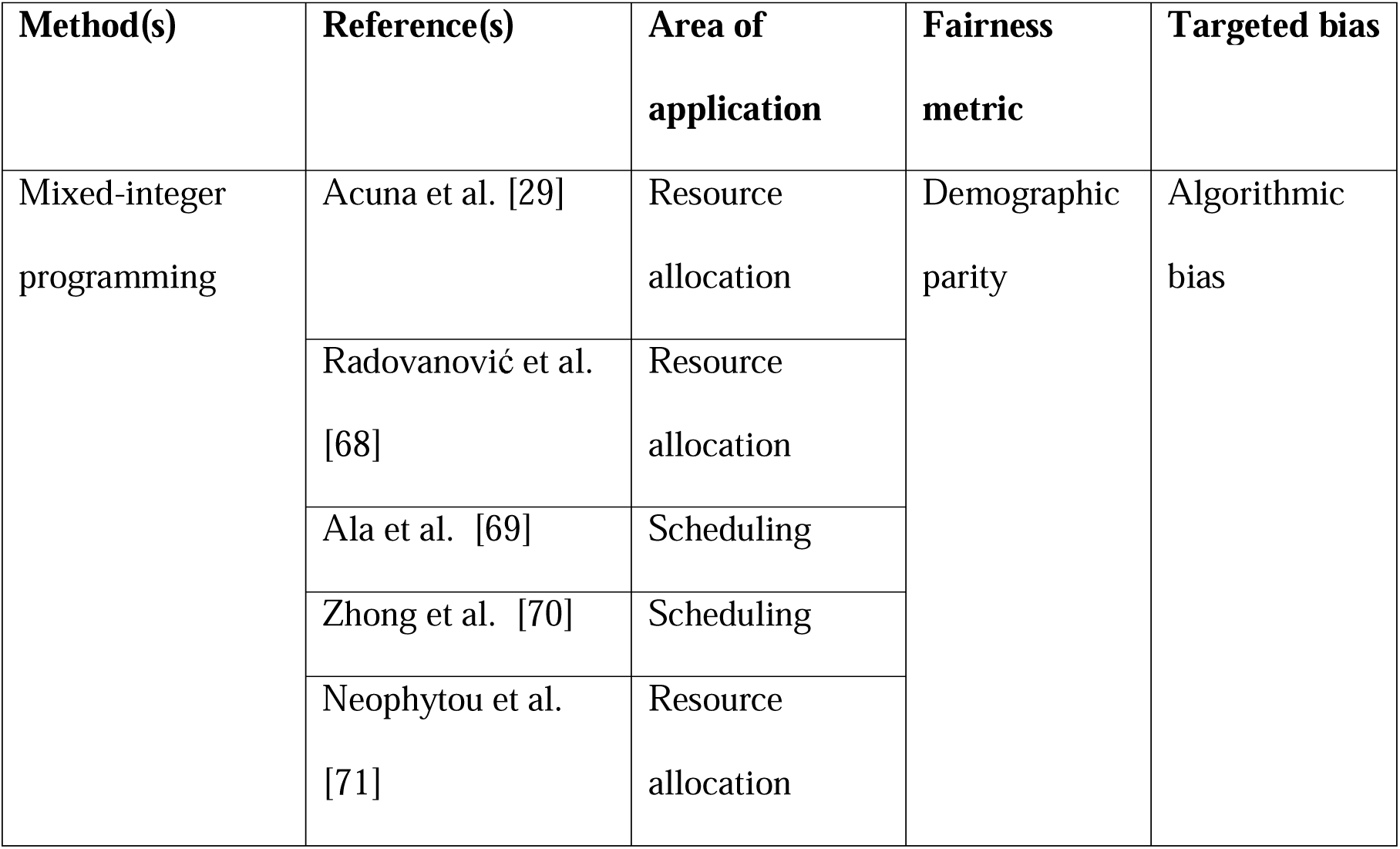
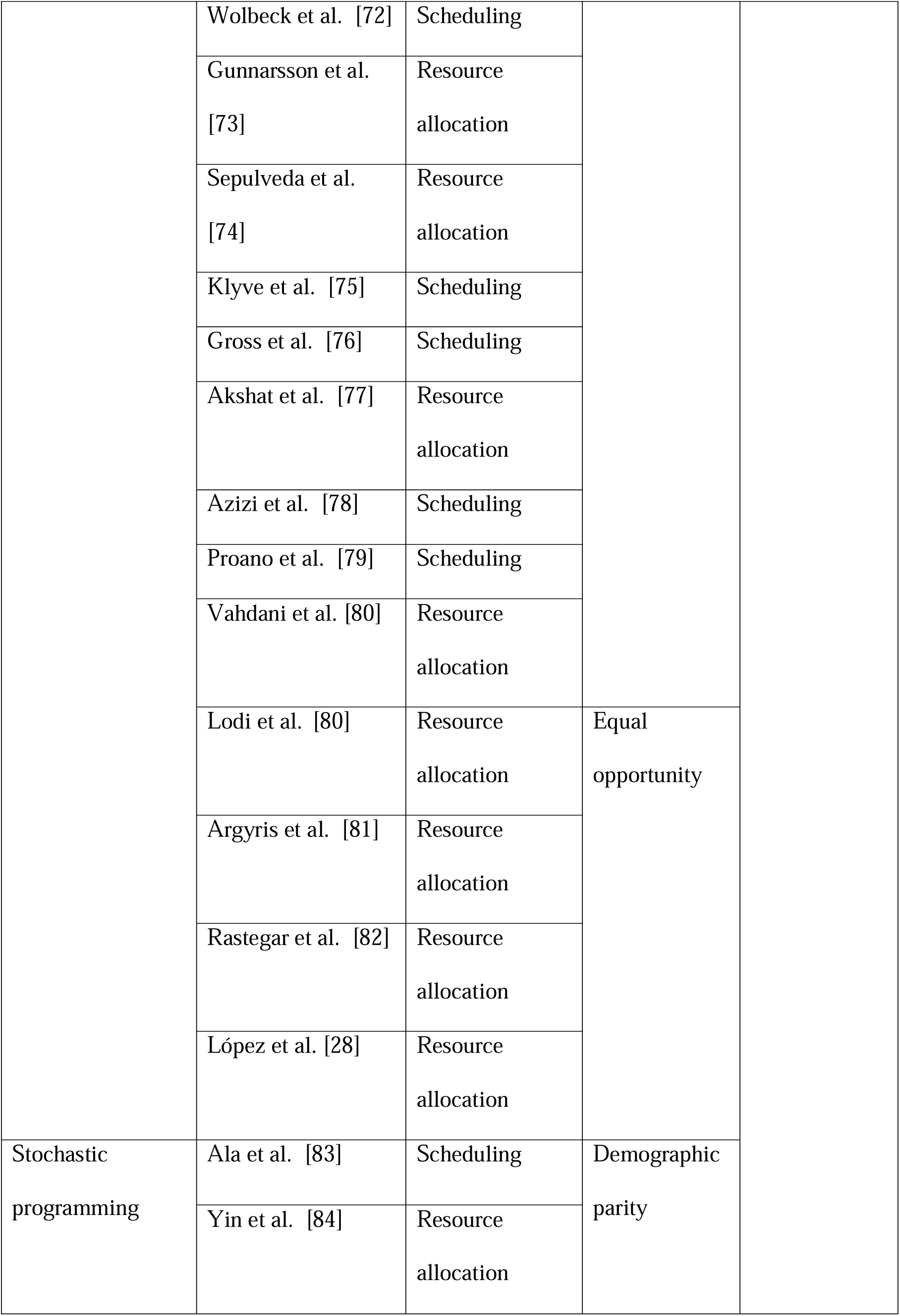
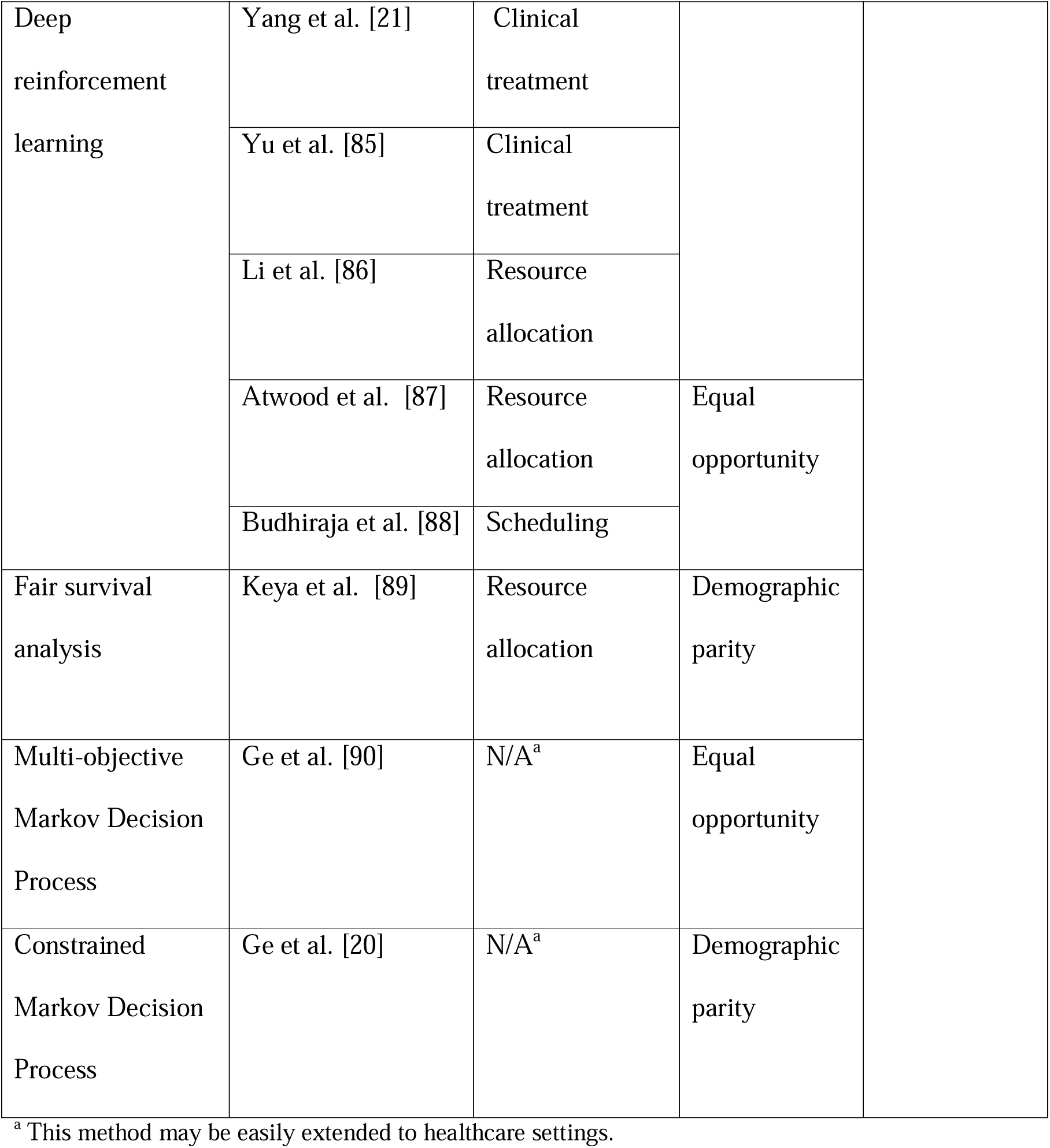
In-processing bias mitigation methods.

| **Method(s)** | **Reference(s)** | **Area of application** | **Fairness metric** | **Targeted bias** |
| --- | --- | --- | --- | --- |
| Mixed-integer programming | Acuna et al. [29] | Resource allocation | Demographic parity | Algorithmic bias |
|  | Radovanović et al. [68] | Resource allocation |  |  |
|  | Ala et al. [69] | Scheduling |  |  |
|  | Zhong et al. [70] | Scheduling |  |  |
|  | Neophytou et al. [71] | Resource allocation |  |  |

|  |  |  |  |
| --- | --- | --- | --- |
|  | Wolbeck et al. [72] | Scheduling |  |
|  | Gunnarsson et al. [73] | Resource allocation |  |
|  | Sepulveda et al. [74] | Resource allocation |  |
|  | Klyve et al. [75] | Scheduling |  |
|  | Gross et al. [76] | Scheduling |  |
|  | Akshat et al. [77] | Resource allocation |  |
|  | Azizi et al. [78] | Scheduling |  |
|  | Proano et al. [79] | Scheduling |  |
|  | Vahdani et al. [80] | Resource allocation |  |
|  | Lodi et al. [80] | Resource allocation | Equal opportunity |
|  | Argyris et al. [81] | Resource allocation |  |
|  | Rastegar et al. [82] | Resource allocation |  |
|  | López et al. [28] | Resource allocation |  |
| Stochastic programming | Ala et al. [83] | Scheduling | Demographic parity |
|  | Yin et al. [84] | Resource allocation |  |

|  |  |  |  |
| --- | --- | --- | --- |
| Deep reinforcement learning | Yang et al. [21] | Clinical treatment |  |
|  | Yu et al. [85] | Clinical treatment |  |
|  | Li et al. [86] | Resource allocation |  |
|  | Atwood et al. [87] | Resource allocation | Equal opportunity |
|  | Budhiraja et al. [88] | Scheduling |  |
| Fair survival analysis | Keya et al. [89] | Resource allocation | Demographic parity |
| Multi-objective Markov Decision Process | Ge et al. [90] | N/A<sup>a</sup> | Equal opportunity |
| Constrained Markov Decision Process | Ge et al. [20] | N/A<sup>a</sup> | Demographic parity |

<sup>a</sup> This method may be easily extended to healthcare settings.

##### Mixed-integer programming (MIP)

This methodology has been widely used to ensure fairness in decision making [29, 68–82]. Emergency department overcrowding has become a nationwide crisis over the last decade [29]. To resolve the overcrowding issue, researchers have applied MIP to build fair medical resource distribution models. An MIP is a type of constrained optimization problem that allows for both integer and continuous variables in its objective and constraints [91]. For example, Acuna and coauthors added equity constraints to ensure that the minimal quality of care for every emergency is greater than or equal to a predefined threshold in an ambulance allocation situation [29]. We discuss this example in subsection 1.2.1 of the Supplementary Information.

Another ubiquitous way to fulfill fairness requirements in decision making is to modify the objective function of an optimization approach. When medical resources are scarce, people from vulnerable groups may have lower access to them. To ensure fairness towards vulnerable populations, the objective function of an algorithm can be set to maximize the smallest number of allocated resources across all population subgroups [28]. This objective ensures that each subgroup receives their required medical support.

##### Stochastic programming

Researchers have also leveraged stochastic programming techniques to generate in-processing bias mitigation techniques [83, 84]. These techniques optimize an objective function while representing uncertainty through probability distributions [92]. For example, we can add fair constraints in stochastic programming models to limit the expected difference between the maximum waiting time and minimum waiting time [83]. Please refer to subsection 1.2.2 of the Supplementary Information for a more detailed discussion of this example.

##### Deep reinforcement learning

Reinforcement learning and deep learning play a pivotal role in in-processing methods. Reinforcement learning is a type of ML where the algorithms learn to make decisions by performing actions and observing the results in an environment of interest [93]. Deep learning applies multiple neural network layers and activation functions to extract new features from the input data [94], being capable of recapitulating and modeling complex patterns in data. Deep reinforcement learning is a combination of deep learning and reinforcement learning that can be used to solve Markov Decision Process (MDP) models. An MDP is a mathematical framework used for modeling decision making in situations where outcomes are partly random and partly under the control of a decision-maker [95]. In an MDP, the reward quantifies how desirable the outcome is after taking a specific action in a given situation. Higher rewards indicate more favorable outcomes. For instance, Yang et. al redefine the rewards of deep reinforcement learning by ensuring the absolute value of rewards of subgroups are smaller if they have a large size [21]. We discuss this example in more detail in subsection 1.2.3 of the Supplementary Information.

##### Fair survival analysis

Fair survival models provide an additional tool for decision making in healthcare settings [89]. Traditional survival analysis estimates the time until an event of interest [96]. Fair survival models incorporate event probabilities and fairness violations when providing the estimate. For example, the allocation sequence of a resource may be decided based on the risk outcomes of a survival model that penalizes the difference between the highest and lowest probabilities of disease incidence within a cohort [89]. Their numerical experiment shows the fair survival model can substantially reduce the group disease risk range. Please refer to subsection 1.2.4 of the Supplementary Information for a more detailed discussion of this example. By integrating these survival-based risk predictions into the waitlist formation, decision makers can systematically balance disease risk considerations with fairness goals when allocating limited resources.

##### Multi-objective Markov Decision Process

While it has not been applied to healthcare settings to the best of our knowledge, the multi-objective MDP is a promising approach to alleviate the potential effect of bias. This model is an extension of the traditional MDP with the difference that the reward function depends on a utility objective and a fairness objective [97]. Ge and coauthors have applied the Pareto frontier to identify the policy that optimizes both utility and fairness elements [90]. Their results show that there exists a trade-off between utility and fairness performance, and we can choose the final policy—i.e., the action chosen in each situation—based on user preferences. We discuss this example in more detail in subsection 1.2.5 of the Supplementary Information. Though multi-objective MDP has not been applied to fair decision making in healthcare to the best of our knowledge, it is possible to deploy these methods to generate fair clinical decisions. For example, if we need to guarantee similar vaccination rates between males and females, we can add this fairness objective into our model. The Pareto frontier can return optimal policies that consider both vaccine utility and distribution fairness.

##### Constrained Markov Decision Process

The Constrained Markov Decision Process (CMDP) is another prospective direction. Compared to traditional MDP, CMDP can accommodate fair constraints in decision making [98]. In CMDP, we can formulate the cost function regarding fairness, and then choose policies leading to fairness cost less or equal to the threshold [20]. This model has not been utilized in fair healthcare decision making to the best of our knowledge. However, we can model fairness metrics as constraints and choose a set of policies that satisfy these constraints. Afterward, we can investigate which policy in this set gives the optimal discounted accumulated reward. Please refer to section 1.2.6 of the Supplementary Information for a more detailed explanation of this example.

##### Summary of In-Processing Methods

Each bias mitigation strategy suits different healthcare scenarios based on data availability, decision objectives, and computational constraints. For instance, mixed-integer programming MIP is effective for discrete resource allocation (e.g., scheduling or capacity planning) and allows explicit fairness constraints, but it may struggle with high-dimensional patient data. Stochastic programming handles uncertainty in clinical pathways and can incorporate probabilistic fairness bounds, yet it requires robust probability estimates and may be computationally demanding. Deep reinforcement learning adapts well to complex state spaces and large-scale data, but it can be opaque, raising explainability concerns and requiring careful regularization to avoid overfitting. Fair survival-based decision-making is crucial for time-to-event outcomes, ensuring that interventions and resources are allocated in a way that balances long-term risks and benefits across demographic groups. However, enforcing fairness at a specific time horizon can introduce temporal distortions in subsequent decisions, as ensuring equity at one point in time may inadvertently shift survival probabilities—and thus decision outcomes—at later points. Multi-objective MDP models allow clinicians to weigh multiple criteria (e.g., treatment effectiveness vs. equity), though specifying trade-offs can be challenging. Constrained MDP models impose hard fairness requirements that are useful for high-stakes decisions, but they may reduce flexibility in optimizing overall performance. Ultimately, the choice depends on the clinical setting, data availability, ethical priorities, and the extent to which one can tolerate reduced performance for stronger fairness guarantees.

#### 3.3.3 Post-Processing

Post-processing methods calibrate algorithmic outcomes to achieve fairness. We identify the following post-processing methods relevant to healthcare applications: Laplacian smoothing, multi-accuracy approaches, and expert systems. The post-processing bias mitigation methods included in this review are shown in Table 4. Please refer to section 1.3 of the Supplementary Information for a more detailed discussion of the strengths and limitations of post-processing methods.

**Table 4:**
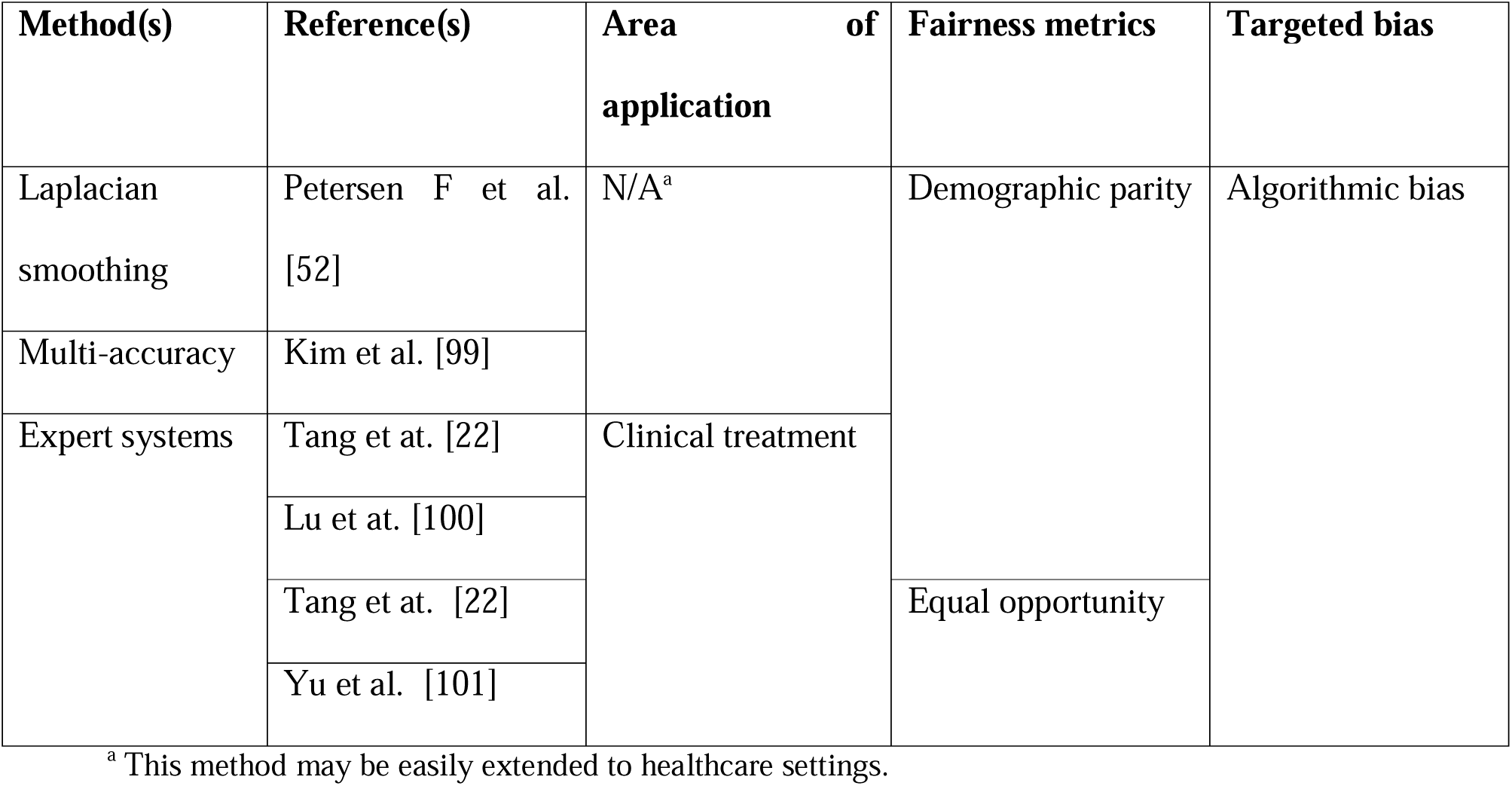
Post-processing bias mitigation methods.

##### Laplacian smoothing

The Laplacian smoothing method is a technique to reduce the noise of the data while preserving the important characteristics of the solution technique [102]. For instance, we can take advantage of this method to guarantee comparable treatment plans among similar individuals while preserving the performance of the algorithms [52]. Selecting an appropriate smoothing parameter is critical: too much smoothing risks diluting meaningful subgroup distinctions, while too little smoothing can leave small groups vulnerable to statistical noise. Balancing these factors is key to leveraging Laplacian smoothing as an efficient and robust tool for improving fairness in ML applications.

##### Multi-accuracy

We can apply multi-accuracy approaches to combine several weak learners to achieve high accuracy rates among all subpopulation groups [99]. These methods play vital role in classification techniques by assigning larger weights to samples identified incorrectly in weak learners. Subsequently, the following weak learners pay extra attention to misidentified samples and adjust their results accordingly [103]. With accurate and fair classification for target populations, physicians can deploy algorithm-based treatment design to achieve desirable medical outcomes.

In addition, the efficiency of multi-accuracy methods hinges on several factors several factors: the complexity and granularity of subgroup identification, the number of iterations required for convergence, and the computational overhead associated with retraining or correcting the model. Efficient implementation demands careful selection of error thresholds to detect performance gaps without incurring excessive retraining cycles.

##### Expert systems

Lastly, the clinical expertise of medical practitioners may help increase the fairness of algorithms [101, 104, 105]. For example, reinforcement learning techniques can suggest several near-equivalent actions, then we can rely on clinicians to decide what actions can lead to the fairest outcome [22]. This approach may enable improved decisions to overcome potential biases while leveraging practitioners’ experience. In practice, one of the most critical factors in expert systems is the complexity of the knowledge base, including the number of rules, the granularity of each rule, and the amount of domain-specific information encoded; as these grow, so does the computational cost of rule matching and inference. Additionally, update frequency—how often new medical guidelines or clinical insights are incorporated—can influence both performance and maintenance overhead.

A summary of the strengths and limitations of the bias mitigation methods presented in this survey is shown in Table 5. We discuss these advantages and disadvantages in more detail in section 1 of the Supplementary Information.

**Table 5:** Strengths and limitations of bias mitigation methods.

| Field | Method(s) | Strengths | Limitations |
| --- | --- | --- | --- |
| Pre-Processing | Reweighting | Preserves original data without generating synthetic samples | Requires careful tuning to avoid overemphasizing minority groups |
|  | Resampling | Addresses class or group imbalance directly | Oversampling can lead to overfitting while undersampling risks losing valuable information |
|  | Fair data transformers | Adjusts or masks sensitive features to reduce bias | May lose important information if transformations are too aggressive |
|  | Natural language processing | Useful for extracting rich features from clinical notes | Computationally intensive for large text corpora |
|  | Post-survey analysis | Provides direct feedback on perceived fairness | Time-consuming to conduct and interpret |
| In-Processing | Mixed-integer programming | Guarantees an optimal solution under well-defined constraints | Computationally expensive for large-scale problems |
|  | Stochastic programming | Handles uncertainty in parameters or data | Modeling complexity increases with more uncertainty sources |
|  | Deep reinforcement learning | Can adapt to changing patient populations over time | Requires large amounts of training data |
|  | Fair survival analysis | Accounts for time-to-event data and censoring | Limited standard tools for enforcing fairness constraints |
|  | Multi-objective Markov Decision Process | Simultaneously optimizes multiple criteria | Finding a balanced solution among competing objectives can be complex |
|  | Constrained Markov Decision Process | Directly enforces fairness or risk constraints within the MDP framework | Potentially high computational overhead for large state/action spaces |
| Post-Processing | Laplacian smoothing | Stabilizes parameter estimates, especially for underrepresented groups | Can over-smooth differences among subgroups |
|  | Multi-accuracy | Ensures balanced performance across subgroups by iterative audits | Repeated audits can be time-consuming and computationally expensive |
|  | Expert systems | Transparent, rule-based reasoning aligned with clinical guidelines | Updating and maintaining rules is labor-intensive, especially for evolving medical knowledge |

The distribution of bias mitigation literature is presented in Supplementary Figure 1 of the section 2 of the Supplementary Information. We include the trend of bias mitigation approaches over the last decade as Figure 3 and the distribution of papers across areas of application in Figure 4.

**Figure 3:**
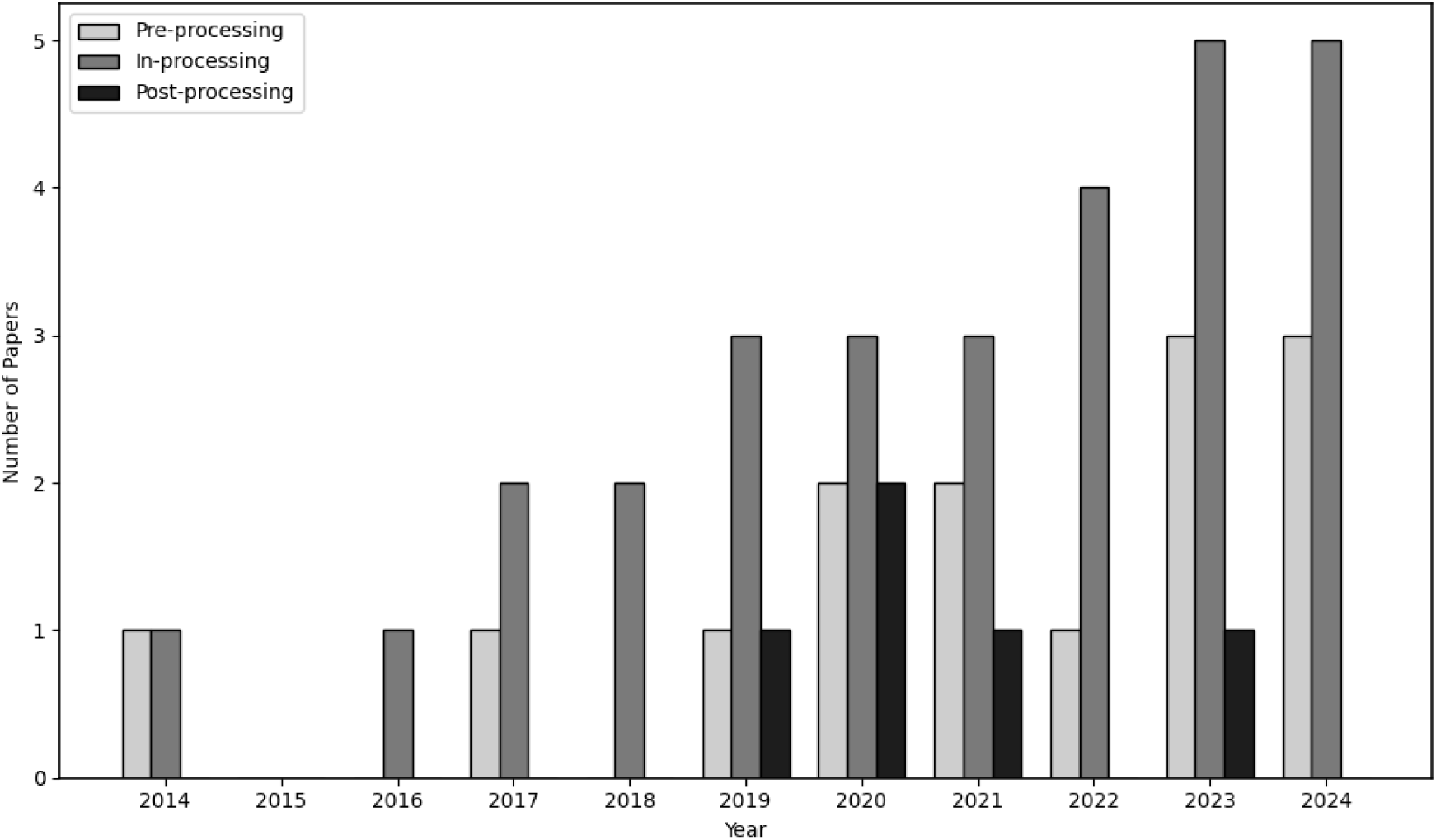
Trend of bias mitigation approaches over the last decade.

**Figure 4:**
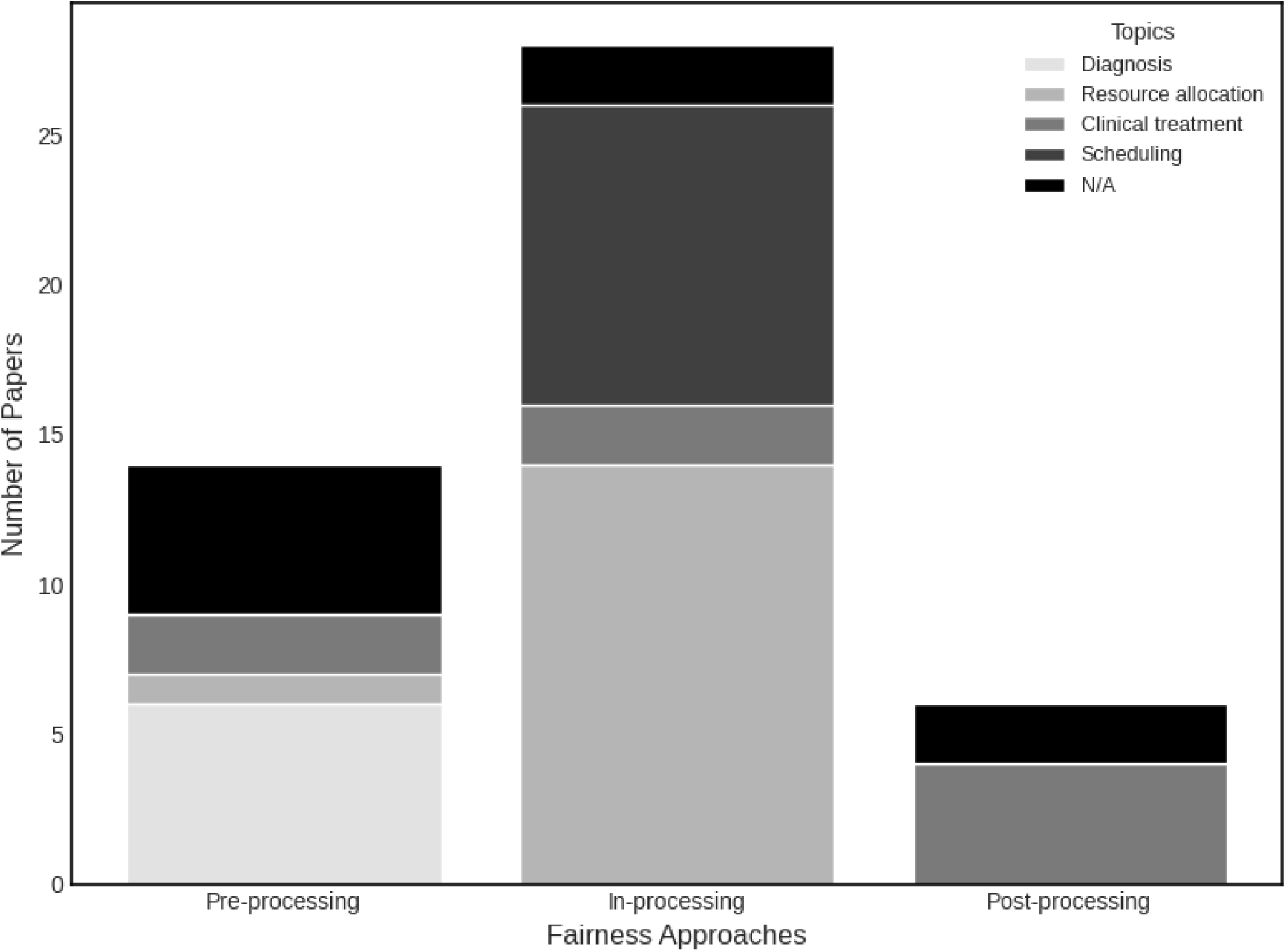
Distribution of papers across topics.

#### 3.3.4 Fair Decision Making Nationally and Internationally

In the U.S., fair decision-making in healthcare is often shaped by regulatory frameworks such as the Health Insurance Portability and Accountability Act (HIPAA), alongside a fragmented healthcare system that influences data-sharing practices [106]. Research in the U.S. tends to focus on algorithmic bias mitigation within electronic health record data, reflecting the prominence of private healthcare providers and diverse data ownership models. Conversely, some non-U.S. settings—especially in countries with nationalized healthcare systems—can leverage centralized patient databases, which may offer more uniform coverage but also raise unique privacy and ethical concerns. Regulatory regimes like the European Union’s General Data Protection Regulation (GDPR) further shape data handling and bias detection by imposing stricter consent and transparency requirements [107]. Moreover, cultural differences in attitudes toward data collection, resource allocation, and equity lead to variations in how fairness metrics are defined and prioritized. Despite these contextual disparities, both U.S. and international studies share common challenges—such as representation bias and response bias—pointing to the need for continued global collaboration to develop robust, context-aware fairness solutions.

## 4. Conclusion and Future Research Directions

Bias in decision making can stem from various factors, such as algorithm design, data sources, publication practices, limited resources, patient diversity, and cultural differences. Commonly discussed fairness metrics include fairness through unawareness, demographic parity, and equal opportunity. Multiple metrics have been developed to conceptualize and quantify fairness across various stages of the decision-making pipeline, including the dataset, algorithmic design, and resulting outcomes. To address biases stemming from these distinct sources, researchers have proposed three primary categories of bias mitigation strategies: (1) *Pre-processing methods*, which focus on identifying and rectifying biases within datasets prior to their use in modeling and algorithms, thereby ensuring more equitable representations; (2) *In-processing methods*, which aim to incorporate fairness constraints or objectives directly into the algorithm and model design, reducing disparities across subgroups during the modeling process; and (3) *Post-processing methods*, which adjust model outcomes to achieve fairer results without altering the underlying algorithm and model. These approaches collectively represent a comprehensive framework for addressing fairness by optimization and ML.

### 4.1 Extensions and Future Work

Continuing research on fair decision making in healthcare is vital because existing methods do not yet fully capture the complexity and diversity of real-world clinical settings. Many current approaches rely on assumptions or metrics that overlook individual patient nuances, such as evolving clinical states or social determinants of health. Moreover, high-stakes healthcare decisions demand both transparency and ethical accountability, though existing “black box” models can obscure sources of bias. As a result, there is a pressing need for new, context-aware frameworks that integrate domain expertise, robust optimization, and interpretability to ensure equitable outcomes for all patients.

Moving forward, several questions are worth exploration: The first interesting open question is explainability for fair algorithms. Many fair algorithms in healthcare are considered black boxes that are hard to understand. The lack of explainability is an obstacle for practitioners to identify if the model is relying on biased features. Another promising field is algorithmic interpretability. If a fair algorithm doesn’t align with practitioners’ intuition and domain knowledge, practitioners may be skeptical to adopt the algorithmic output. In addition, it is worthwhile to explore how to keep the balance between interpretability and fairness. While increased interpretability can earn more trust among practitioners, it may hurt the model’s fairness [64]. Furthermore, context-aware fairness metrics is a significant topic to explore. Researchers have found that different fairness metrics can be incompatible. Thus, we cannot expect a model to satisfy all fairness metrics [103]. In such contexts, it is critical to understand which type of metric we should consider in a specific circumstance. Identifying the best fairness metric for a specific problem will likely require cooperation between modelers and domain experts [26]. Another promising methodological innovation is to bridge the divide between fair prediction and fair decision making. Typically, current approaches follow a “fair prediction then optimization” pipeline, operating under the assumption that fair predictions will naturally result in fair decisions. However, this causal link is not assured. To address this divide, innovative methodologies are required that explicitly incorporate the cost of fair decision making—stemming from fair predictions—into the optimization objective. Such approaches could enable the simultaneous achievement of both fair prediction and fair optimization [108].

Furthermore, we need to consider fairness beyond expected values, and extensions such as fairness of variances across subgroups can fulfill the goal by evaluating the spread of outcomes, not just their expectations [21]. In addition, ideas from unsupervised and supervised learning, such as representation learning, can reveal hidden subgroups for personalized care before policy optimization, supporting fair decision making in healthcare [55]. Finally, another big open question is to establish individual-level fairness in large populations. Individual-level fairness is the notion that two individuals who are sufficiently similar in clinically relevant ways should receive similar outcomes. Establishing this notion within large and diverse healthcare populations remains an open challenge. This task involves defining appropriate measures of similarity in complex, high-dimensional data and ensuring that these measures do not conflict with broader group-level fairness criteria or clinical objectives. Successfully addressing individual-level fairness would enable more personalized and equitable decision making, ultimately improving patient care [49].

We acknowledge that factors such as information asymmetry, resource constraints, patient diversity, and cultural differences can intersect with or exacerbate the biases discussed in this review. For instance, limited or skewed data can intensify representation bias, and uneven access to resources may deepen representation bias. Although a detailed exploration of these broader challenges is beyond our current scope, we note their significance and encourage future research to address them in conjunction with the biases we identify. Furthermore, the bias mitigation techniques mentioned in subsection 3.3 can be applied to various healthcare challenges, including information asymmetry, limited resources, patient diversity, and cultural differences. For example, reweighting or resampling can address data imbalances from vulnerable groups, while fairness constraints may help ensure each group receives a minimum standard of care. Furthermore, methods like constrained MDP models can dynamically adapt recommendations to diverse patient characteristics, promoting more equitable outcomes.

Because our search was restricted to English-language sources, this review may not fully capture the perspectives and developments present in non-English-speaking regions. Additionally, we might have missed keywords during our literature review, leading to the omission of works that used excluded terms.

### 4.2 Practical Considerations

The main sources of agreement and disagreement in fair decision-making in healthcare revolve around fairness metrics, algorithmic approaches, and their practical deployment. There is broad agreement on the importance of addressing biases in healthcare data and decision-making processes, with researchers emphasizing the necessity of fairness-aware methodologies such as preprocessing, in-processing, and post-processing techniques. However, disagreements arise in the selection and application of fairness metrics, as different metrics often produce conflicting results [103]. Moreover, there is disagreement over the assumption that fair predictions automatically lead to fair decision making, highlighting the need for more integrated approaches [105]. These points of divergence underscore the complexity of developing fair healthcare methodologies and the necessity for context-sensitive, interdisciplinary solutions [94].

To effectively deploy methods above, practitioners need accessible tools and frameworks that bridge the gap between theoretical advancements and real-world applications. First, the development of user-friendly software libraries and platforms incorporating fairness-aware algorithms and explainability features can empower practitioners to implement these methods without requiring extensive technical expertise. Open-source fair ML libraries, such as Fairlearn in Python and fairml in R, provide a valuable foundation for implementing bias mitigation strategies in predictive modeling, yet ongoing commitments are necessary to address the full complexity of implementing fair decision-making methods. Second, integrating domain-specific knowledge into these tools is critical, as it enables models to be tailored to the unique challenges and priorities of healthcare. Third, practitioners should engage in interdisciplinary collaboration with data scientists, ethicists, and domain experts to ensure that fairness and interpretability align with practical needs and ethical standards. Finally, regular audits and monitoring systems should be established to continuously evaluate the performance and fairness of deployed models, providing feedback loops to refine methodologies and address emerging biases. By adopting these approaches, practitioners can effectively leverage innovative fairness methods to create decision-making systems that are robust, equitable, and transparent.

For researchers, our work offers a comprehensive synthesis of state-of-the-art fairness approaches, highlighting current trends, identifying research gaps, and suggesting promising future directions. This review not only facilitates methodological comparisons and benchmarking but also serves as a roadmap for developing novel fairness metrics, mitigation strategies, and evaluation frameworks. By integrating insights from both theory and practice, our survey aims to help researchers advance the field and stimulate further research into robust, equitable, and transparent decision-making systems in healthcare.

### 4.3 Conclusions

Given the growing importance of decision-making approaches in healthcare, fairness considerations and bias-mitigation approaches are increasingly vital. Our survey may aid practitioners and researchers in 1) understanding potential sources of biases in decision making; 2) choosing the appropriate fairness metric to evaluate decision-making models; and 3) selecting the appropriate pre-processing, in-processing, and post-processing techniques to reduce bias. In conclusion, this survey sheds light on the current state and challenges of fair decision-making in healthcare, highlighting the crucial need for continuous improvement in policies and practices to ensure equitable healthcare outcomes for all individuals.

## Supporting information

Supplementary information on pre-processing, in-processing, and post-processing methods

## Data Availability

All data produced are available online at Google Scholar.

https://scholar.google.com/

## Statements and Declarations

## Acknowledgments

The financial support for this study was provided in part by the Thayer Tuition Fellowship at Dartmouth College. The funding agreement ensured the authors’ independence in designing the study, interpreting the data, writing, and publishing the report.

