## Supplementary information on pre-processing, in-processing, and post-processing methods for "A Survey on Optimization and Machine Learning-Based Fair Decision Making in Healthcare"

### 1. Bias Mitigation Techniques Details

#### 1.1 Pre-Processing Methods

##### 1.1.1 Reweighting

Strengths of Reweighting

Reweighting methods are most appropriate in situations where underrepresented groups are clearly identifiable, and the disparity in representation is a primary driver of model bias. For instance, in medical studies where specific racial groups are underrepresented in genomic data, reweighting can help ensure that the model treats these groups fairly by giving their data more influence during training.

Limitations of Reweighting

Reweighting methods rely on accurate estimates of population weights or probabilities, and any inaccuracies in these estimates can skew the model further, leading to unstable or unintended outcomes [1]. Moreover, excessively large weights for underrepresented instances can result in overfitting, where the model becomes overly sensitive to small variations in the weighted data, potentially degrading performance on the majority population or leading to fairness-performance trade-offs. These challenges highlight the need for careful implementation and validation of reweighting strategies to ensure that they genuinely reduce bias and do not inadvertently create new disparities.

##### 1.1.2 Resampling

Strengths of Resampling

Resampling is particularly beneficial in scenarios where data imbalance is the primary source of bias, and the minority group instances are too sparse for the model to learn meaningful patterns. For example, Chawla and collaborators deploy a synthetic minority resampling technique to decide whether a patient needs diabetes treatment. Their method successfully shrinks the gap of true positive rates between majority and minority groups [2].

Limitations of Resampling

The relative proportions of protected groups heavily influence efficiency of resampling. Medical data are typically complex, and resampling may lead to overfitting because the model memorizes patterns in the resampled data rather than generalizing to unseen cases [3]. In the context of diabetes treatment, if undersampling the majority group removes meaningful instances that represent real-world scenarios, the model might fail to generalize effectively to the entire population [4].

##### 1.1.3 Fair Data Transformers

Strengths of Fair Data Transformers

Fair Data Transformers leverage the foundational strengths of transformer models—such as their ability to model complex, sequential, or high-dimensional data—while embedding fairness constraints into the learning process. The transformer can capture nuanced patterns without perpetuating biases in downstream decision making [5]. In practice, these mechanisms ensure that the model maintains near-optimal decision-making efficacy while systematically reducing bias, thereby supporting ethical outcomes in sensitive healthcare applications.

Limitations of Fair Data Transformers

Researchers have observed that among data transformers, selecting a subset of features can introduce unfairness. In addition, feature standardization and non-linear transformers can be biased under special conditions such as having too many outliers. These observations indicate that the appropriate transformer must be selected on a case-by-case basis. Furthermore, transformers such as principal component analysis may prioritize fairness but could reduce interpretability by transforming features into latent dimensions, making it harder to justify decisions to stakeholders [5].

##### 1.1.4 Natural Language Processing

Strengths of Natural Language Processing

One of the key advantages of natural language processing is its ability to handle large, unstructured text datasets, such as clinical notes, where sensitive information may be deeply embedded and difficult to isolate manually. Natural language processing ensures the algorithms do not consider sensitive attributes during decision making. For example, Minot and coauthors identify and remove gender-related languages from electronic health records by using bidirectional encoder representations. Then, they deploy classification algorithms to evaluate health conditions and give clinical suggestions. Their results show that fairness across genders improves with only a mild degradation in performance [6]. In addition, natural language processing can be trained on multilingual and culturally diverse datasets, enhancing their ability to accurately interpret symptom descriptions from non-native speakers and capture subtle linguistic nuances [7, 8].

Limitations of Natural Language Processing

Removing sensitive information entirely may lead to a loss of context or critical information, potentially reducing the efficacy of downstream decision-making algorithms. For instance, while removing gendered language may improve fairness, it could inadvertently strip out clinically relevant details that are correlated with gender but essential for diagnosis or treatment (e.g., gender-specific symptoms). Natural language processing also has substantial computational cost, which can significantly influence the processing efficiency. For example, the number and complexity of preprocessing steps (e.g., tokenization, stemming, lemmatization) can significantly increase runtime, especially for large-scale or real-time healthcare applications [7]. Moreover, large language models  are computationally intensive to train and fine-tune, which can strain memory and processing resources [9].

##### 1.1.5 Post-survey Analysis

Strengths of Post-survey Analysis

Post-survey analysis can identify and address gaps in the data—such as missing values and outliers—to enhance data quality. By refining the underlying information in this way, it reduces disparities across different groups or stakeholders, ensuring subsequent decisions are based on more complete and equitable evidence.

Limitations of Post-survey Analysis

Post-survey analysis is highly sensitive to the robustness of the statistical methods used to analyze subgroup responses: techniques that fail to account for intersecting protected characteristics (e.g., race and gender simultaneously) may overlook significant disparities. Additionally, post-survey adjustments often rely on assumptions or statistical models that may not fully capture the complexity of the underlying biases, potentially leading to overcorrection or undercorrection [10].

#### 1.2 In-Processing Methods

##### 1.2.1 Mixed-Integer Programming (MIP)

Example use of MIP

Acuna and coauthors added equity constraints to their MIP formulation to ensure that the minimal quality of care for every emergency is greater than or equal to a threshold $\beta$ in an ambulance allocation problem [11]. The equity constraints in their MIP guarantee patients suffering from uncommon diseases still receive necessary clinical support. The equity constraints are demonstrated below:

$$\sum_{j \in J} q_{\{i,j\}} X_{\{i,j\}} \geq \beta, \ \forall i \in I,$$

where $I$ denotes the set of all possible diseases and $i \in I$ denotes disease $i$. Moreover, $J$ denotes the set of all emergency departments, $j \in J$ refers to the emergency department $j$, $q_{\{i,j\}}$ is the quality of care for disease $i$ offered by emergency department $j$, and $X_{\{i,j\}}$ is a binary decision variable. If department $j$ provides the care for disease $i$, then $X_{\{i,j\}}$ =1, otherwise $X_{\{i,j\}} = 0$. Lastly, $\beta \in [0,1]$ is selected based on domain experts' suggestions, where 0 denotes the worst quality and 1 the best quality.

Strengths of MIP

Mixed-integer programs excel in fair healthcare decision making by providing a structured framework that precisely formulates complex decision-making problems with explicit objectives or built-in fairness constraints. Such mechanism can enhance the transparency and explainability of the model.

Limitations of MIP

The limitation of MIP lies in scalability. When the problem size grows exponentially with the number of constraints and decision variables, MIP can be computationally expensive for large-scale or real-time scenarios, such as nationwide vaccine distribution.

##### 1.2.2 Stochastic Programming

Example use of Stochastic Programming

To optimize patients' waiting time, we can add fair constraints in stochastic programming models to limit the expected difference between the maximum waiting time and minimum waiting time across patients. The constraint can be formularized as [12]:

$$\max_{n,k} \mathrm{E}\left(W_{\{k\}}^{\{n\}}\right) - \min_{n,k} \mathrm{E}\left(W_{\{k\}}^{\{n\}}\right) \leq \alpha.$$

Here, $k = 1,2, \ldots T$ denotes the time slots when decisions are made, $n$ denotes the $n$-th patient, and $\alpha \geq 0$ is the threshold suggested by domain experts. The expected waiting time of the $n$-th patient scheduled to interval $k$ is represented by $E\left(W_{\{k\}}^{\{n\}}\right)$. These constraints guarantee the expected waiting time among all patients does not vary drastically.

Strengths of Stochastic Programming

Stochastic programming's primary strength in fair decision-making is its ability to systematically incorporate uncertainty into optimization models. This approach allows decision-makers to plan for multiple scenarios—such as variations in patient demand, resource availability, and treatment outcomes—while embedding fairness constraints directly into the model.

Limitations of Stochastic Programming

The effectiveness of stochastic programming depends heavily on the accuracy of the probabilistic models used to represent uncertainty. These models are typically built using historical data or expert predictions, which may not always fully capture the true variability or complexity of future events. Poor estimates can lead to suboptimal or biased outcomes, particularly in healthcare, where inaccuracies can exacerbate existing inequities. For instance, if future infection rates in underserved regions are underestimated, the optimization model might allocate fewer resources, such as ventilators or vaccines, to those regions. This under-allocation could worsen health disparities, as the model would effectively prioritize regions with overestimated demand.

##### 1.2.3 Deep Reinforcement Learning

Example use of Deep Reinforcement Learning

Yang et. al redefine the rewards of deep reinforcement learning to achieve fairness [13]. In their approach, the absolute value of rewards of a certain subgroup are smaller if the size of the group is large. The reward function is demonstrated below:

$$R(s_t, a_p, l_p) = \begin{cases} \lambda_p, & if\ a_p = l_p \\ -\lambda_p, & if\ a_p \neq l_p \end{cases}.$$

Here, $s_t$ denotes the state at time $t$, $a_p$ denotes the diagnosis of the model for a person in group $p$, and $l_p$ denotes the ground truth disease of the patient from group $p$. The parameter $\lambda_p$ is the reward of group $p$ adjusted by its size. Specifically, a positive reward is given if the agent makes the correct diagnosis, and a negative reward is given otherwise. The authors require the absolute reward for minorities to be greater than the absolute reward of majorities. This definition of rewards helps the solution approach give more attention to minority groups.

Strengths of Deep Reinforcement Learning

In practice, an MDP may encompass a massive number of system configurations (i.e., states), becoming computationally intractable by traditional reinforcement learning methods (e.g., Q learning) [14]. Deep reinforcement learning can take advantage of deep learning to represent a policy (i.e., sequence of procedures for decision making at each state) as a neural network and learn to find a policy that optimizes model outcomes (i.e., rewards). Therefore, deep reinforcement learning can accommodate large or continuous state and action spaces, which Q-learning cannot, given that Q-learning relies on simple Q-tables to generate policies.

Limitations of Deep Reinforcement Learning

One major challenge of deep reinforcement learning is its lack of explainability, as deep reinforcement learning models are often treated as "black boxes," making it difficult for healthcare practitioners to understand or trust the rationale behind decisions, especially in fairness-sensitive applications like resource allocation. In addition, deep reinforcement learning models are computationally intensive, making them challenging to deploy in real-time or resource-constrained settings such as rural hospitals or emergency scenarios [15].

##### 1.2.4 Fair Survival Analysis

<u>Example use of Fair Survival Analysis</u>

Keya et. al remodel the loss function of survival analysis to achieve fairness [16]. The objective of the fair model is below:

$$g(\beta) = -\left(L_{x(\beta)} + \lambda F_{x(\beta)}\right),$$

where $L_{x(\beta)}$ is the log-likelihood of a Cox proportional-hazards model that measures the probability of getting a disease during a certain period and $F_{x(\beta)}$ is the fairness penalty. Moreover, $\lambda$ is the weight of the fairness penalty in the objective. The difference between the highest and lowest probabilities of disease incidence within a cohort is utilized as the metric for evaluating fairness. Then, they feed the input data to train the model (i.e., learn the parameters $\beta$ to optimize the objective). The outcome is used to generate a waitlist of patients, which decides the sequence of resource allocation. Their numerical experiment shows the fair survival model can substantially boost the group disease risk range.

<u>Strengths of Fair Survival Analysis</u>

Fair survival analysis handles censored data and time-to-event outcomes to estimate the likelihood of a specific event occurring within a given timeframe. By leveraging this probabilistic approach, it facilitates more equitable and informed decision making in environments characterized by uncertainty.

Limitations of Fair Survival Analysis

Implementing fair survival analysis often requires rich, representative datasets across all relevant subgroups. In cases of imbalanced or sparse data, especially for minority populations, the fairness constraints may lead to unstable estimates or reduced model robustness [17].

##### 1.2.5 Multi-objective Markov Decision Process

Example use of Multi-objective Markov Decision Process

Ge et. al modify the reward function in Markov Decision Process to ensure fairness in decision making [18]. The modified reward function is:

$$f_w\big(R(s,a)\big) = w^T R(s,a),$$

where $R(s,a)$ is a reward vector containing rewards $r$ for all objectives after taking the action $a$ at state $s$, and $w$ is the weight for each objective. The authors apply reinforcement learning to learn the weight $w$.

Strengths of Multi-objective Markov Decision Process

Multi-objective Markov Decision Process provides a structured and explicit manner to simultaneously optimize multiple, potentially conflicting objectives—such as patient outcomes, resource efficiency, and fairness—in healthcare. This holistic approach ensures that decision-making in complex healthcare environments remains both efficient and equitable for diverse patient populations.

Limitations of Multi-objective Markov Decision Process

The first limitation of multi-objective MDP is the need for precise quantification and prioritization of objectives. In healthcare, it can be difficult to assign appropriate weights to fairness versus clinical outcomes, as these trade-offs often involve subjective or ethical considerations. Furthermore, multi-objective Markov Decision Process models may generate Pareto-optimal policies that require decision-makers to manually select a policy from a set of non-dominated solutions, which can be impractical without clear guidance.

##### 1.2.6 Constrained Markov Decision Process (CMDP)

Example use of CMDP

Ge et. al incorporate a fairness constraint into Markov Decision Process to limit the fairness violation [19]. The constraint can be formulated as:

$$E\left[\sum_{t=0}^{\infty} \gamma\, C_t\right] \leq d,$$

where $C_t$ denotes the fairness cost at time $t$, $\gamma \in (0,1)$ is a discounted factor representing the fairness violations at the current time over the future, and $d$ denotes the threshold for accumulated discounted fairness cost.

Strengths of CMDP

While standard MDP focuses purely on maximizing expected rewards, CMDP extends this framework by embedding fairness or ethical constraints directly into the decision-making process, offering a more robust solution compared to traditional MDPs. Additionally, CMDPs can produce a series of fair decisions over time, ultimately offering long-term benefits to all stakeholders.

Limitations of CMDP

However, CMDPs are less suitable in highly dynamic or uncertain settings, such as during a pandemic, where fairness constraints or system dynamics may evolve rapidly. In a pandemic, factors like infection rates, resource availability, or the vulnerability of different groups can change frequently, making it difficult to define static fairness constraints [20].

#### 1.3 Post-Processing Methods

##### 1.3.1 Laplacian Smoothing

Strengths of Laplacian Smoothing Method

Researchers have shown this technique may improve outcome consistency significantly [21]. Although not applied in healthcare settings yet, the Laplacian smoothing method can be extended to this domain. For example, after a reinforcement learning algorithm produces treatment plans, a Laplacian smoothing method can guarantee comparable treatment plans are assigned to patients with similar severeness, regardless of patients' sensitive attribute.

Limitations of Laplacian Smoothing Method

Laplacian smoothing can inadvertently oversmooth the data or decisions, erasing meaningful differences. For example, if two patients have similar blood pressure levels but different underlying conditions (e.g., one has diabetes while the other does not), oversmoothing might assign both the same treatment plan, which could be harmful for one of them.

##### 1.3.2 Multi-accuracy Approaches

Strengths of Multi-accuracy Approaches

Multi-accuracy approaches can significantly improve the accuracy rate of subgroups with the worst classification error [22], which shows a promising future for complex problems such as population health assessment.

<u>Limitations of Multi-accuracy Approaches</u>

However, overfitting can occur when the multi-accuracy approaches overly emphasize correcting errors for minority subgroups, potentially degrading performance for the overall population. In addition, the efficiency of multi-accuracy methods hinges on several factors several factors: the complexity and granularity of subgroup identification, the number of iterations required for convergence, and the computational overhead associated with retraining or correcting the model. Efficient implementation demands careful selection of error thresholds to detect performance gaps without incurring excessive retraining cycles.

##### 1.3.3 Expert Systems

<u>Strengths of Expert Systems</u>

This approach can facilitate more informed decisions by leveraging practitioners' expertise and mitigating potential biases [15]. In addition, it maintains transparency through a rule-based reasoning structure that aligns with established clinical guidelines.

<u>Limitations of Expert Systems</u>

The limitation of expert systems is that decisions may be influenced by clinicians' implicit biases, leading to inequities, and the lack of consistency among experts can result in varying outcomes for similar cases. The efficiency of an expert system in clinical decision making is shaped by the complexity of the knowledge base, including the number of rules, the granularity of each rule, and the amount of domain-specific information encoded. As these factors grow, so does the computational cost of rule matching and inference.

### 2. Distribution of Papers Across Bias Mitigation Approaches

The distribution of bias mitigation techniques across the identified papers is demonstrated in Supplementary Figure 1.

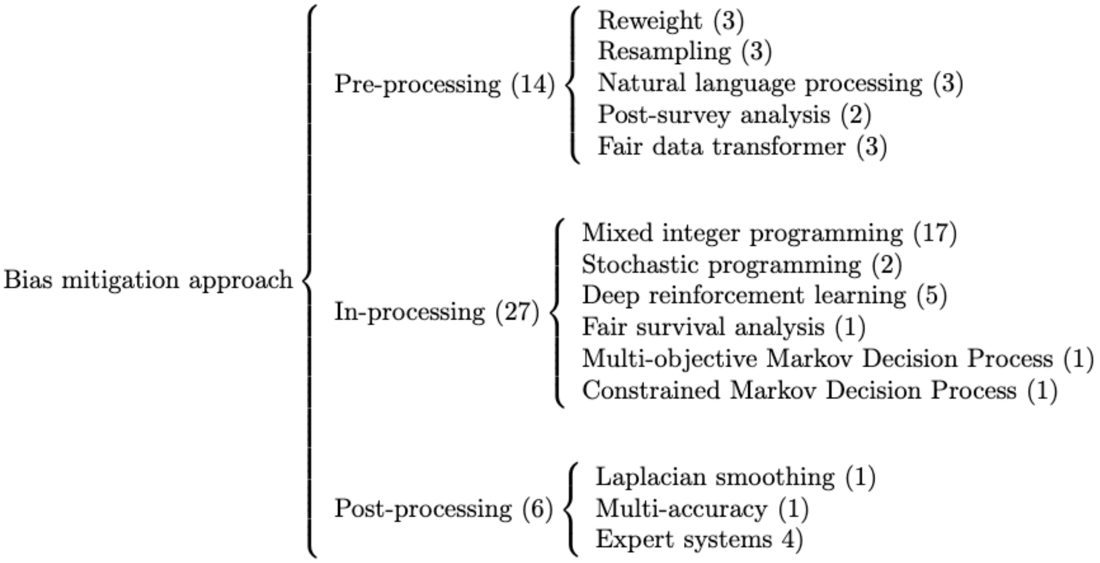


Supplementary Figure 1: Distribution of papers in bias mitigation approach
